# Altered neutrophil phenotype and function in non-ICU hospitalised COVID-19 patients correlated with disease severity

**DOI:** 10.1101/2021.06.08.21258535

**Authors:** KBR Belchamber, OS Thein, J Hazeldine, FS Grudzinska, MJ Hughes, AE Jasper, KP Yip, E Sapey, D Parekh, DR Thickett, A Scott

**Author notes:** **Corresponding author**: Professor David Thickett, Institute of Inflammation and Ageing, University of Birmingham, Birmingham B15 2TH, UK. joint first author (KBR Belchamber, OS Thein). joint senior author (DR Thickett, A Scott). **Author contributions**: Concept and design: KBRB, OST, JH, ES, DP, DRT, AS. Neutrophil work: KBRB, OST. Patient recruitment: OST, FSG, KPY. Additional laboratory work: MJH, AEJ, AS. Drafting manuscript: KBRB, OST, ES, DP, DRT, AS. All authors have read and approved the manuscript. This article has an online supplement. **Sources of funding**: OST/DT/DP; Birmingham Health partners, KBRB/ES; Alpha One Foundation (617303), JH; National Institute for Health Research (NIHR) Surgical Reconstruction and Microbiology Research Centre, ES/KYP; British Lung Foundation (PPRG16), DT/AS/ES/FSG; Dunhill trust (RTF1906\86), ES; Health Data Research-UK, DT/DP; Health Technology Assessment, DT/AS/ES; Health Technology Assessment (NIHR129593), DT/AS/ES; Medical Research Council (MR/L002736/1).

## Abstract

**Rational:** Infection with the SARS-CoV2 virus is associated with elevated neutrophil counts. Evidence of neutrophil dysfunction in COVID-19 is based predominantly on transcriptomics or single functional assays. Cell functions are interwoven pathways, and so understanding the effect of COVID-19 across the spectrum of neutrophil function may identify tractable therapeutic targets.

**Objectives:** Examine neutrophil phenotype and functional capacity in COVID-19 patients versus age-matched controls (AMC)

**Methods:** Isolated neutrophils from 41 hospitalised, non-ICU COVID-19 patients and 23 AMC underwent *ex vivo* analyses for migration, bacterial phagocytosis, ROS generation, NET formation (NETosis) and cell surface receptor expression. DNAse 1 activity was measured, alongside circulating levels of cfDNA, MPO, VEGF, IL-6 and sTNFRI. All measurements were correlated to clinical outcome. Serial sampling on day 3-5 post hospitalisation were also measured.

**Results:** Compared to AMC, COVID-19 neutrophils demonstrated elevated transmigration (p=0.0397) and NETosis (p=0.0366), but impaired phagocytosis (p=0.0236) associated with impaired ROS generation (p<0.0001). Surface expression of CD54 (p<0.0001) and CD11c (p=0.0008) was significantly increased and CD11b significantly decreased (p=0.0229) on COVID-19 patient neutrophils. COVID-19 patients showed increased systemic markers of NETosis including increased cfDNA (p=0.0153) and impaired DNAse activity (p<0.0.001). MPO (p<0.0001), VEGF (p<0.0001), TNFRI (p<0.0001) and IL-6 (p=0.009) were elevated in COVID-19, which positively correlated with disease severity by 4C score.

**Conclusion:** COVID-19 is associated with neutrophil dysfunction across all main effector functions, with altered phenotype, elevated migration, impaired antimicrobial responses and elevated NETosis. These changes represent a clear mechanism for tissue damage and highlight that targeting neutrophil function may help modulate COVID-19 severity.

## Introduction

Coronavirus disease 2019 (COVID-19), caused by the severe acute respiratory syndrome coronavirus 2 (SARS-CoV2) virus, was declared a global pandemic by The World Health Organisation on 11^th^ March 2020 [1]. Up to 80% of people infected with SARS-CoV2 experience mild to moderate respiratory disease, but in 10-20% of cases, infection can manifest as pneumonitis, with 5% progressing to acute respiratory distress syndrome (ARDS). The overall worldwide mortality rate is 2.2% but this increases to up to 50% in the presence of ARDS [2].

Dysregulated virus induced host-immune responses are thought to be the primary cause of severe COVID-19 [3]. Neutrophils are frontline effector cells that protect against rapidly dividing pathogens and play a pivotal role in the antiviral immune response [4]. Early during an inflammatory event, neutrophils migrate into lung tissue where they perform phagocytosis and release proteases, reactive oxygen species (ROS) and as a later response, neutrophil extracellular traps (NETs) to aid clearance of infection [5].

Advanced age is a recognised risk factor for severe COVID-19, including the development of ARDS [6-8]. Ageing is associated with changes in neutrophil function, including reduced migratory accuracy [9], phagocytosis [10] and NETosis [11], which may delay pathogen clearance and increase bystander tissue injury. Further impairments in neutrophil function have been observed in patients with pneumonia [12], sepsis [13] and ARDS [14]. This dysfunction is most apparent in older adults with severe infections [15] and is associated with poor clinical outcomes [12, 13]. Importantly, these functional deficits appear amenable to therapeutic correction, especially in early or less severe infective episodes, suggesting that neutrophils, or their products, may form a tractable target in inflammatory disease [16, 17]. There is some evidence that neutrophil dysfunction could be associated with distinct cellular phenotypes within the neutrophil population, with immature, senescent, activated and pro-inflammatory neutrophils identified in age and disease [13, 18, 19].

Emerging studies suggest that neutrophils are implicated in the pathogenesis of severe COVID-19 and every reporting study has described cell dysfunction (or inferred cell dysfunction through transcriptomics) which could contribute to tissue damage and secondary infection [20-27] However, studies have often been small, fail to provide age-matched controls, assess patients on the intensive care unit (ICU) after a considerable delay since symptom onset, and often consider neutrophil functions in isolation.

These are important considerations. As neutrophil functions change with age, age matched controls are important to identify pathological differences. Although approximately 12% of hospitalised COVID-19 require ICU support, the majority of hospital admissions and deaths occur on non-ICU wards [28]; understanding how to improve outcomes for this patient group is vital. Most studies in sepsis have demonstrated cell function is more therapeutically tractable during early disease (before ICU care is needed), so the timeliness of intervention is important. Cell functions are enabled by interwoven cell pathways with important differences in internal signalling; knowing which functions are impaired informs which therapeutic strategies might restore/maintain all facets of cellular function.

We hypothesised that neutrophils from COVID-19 patients would exhibit diverse altered effector functions and changes to phenotype, with the degree of dysfunction associated with adverse patient outcomes.

This study aimed to perform, for the first time, a comprehensive assessment of *ex vivo* neutrophil phenotype and function in a statistically powered cohort of hospitalised, non-ICU SARS-CoV2 infected patients compared to aged-matched controls and investigate relationships with clinical outcomes.

## Methods

### Healthy donor and patient recruitment

Recruitment is summarised in Figure 1. COVID-19 patients were recruited January to March 2021 from the Queen Elizabeth Hospital Birmingham, in accordance with ethics REC ref: 19/WA/0299 and 20/WA/0092 approved by the West Midlands – Solihull research ethics committee. Written informed consent was obtained where possible; patients unable to consent due to lack of capacity were consented by proxy; designated consultee via telephone or professional consultee. Follow-up samples were collected at days 3-5 with confirmed consent, where possible.

**Figure 1.**
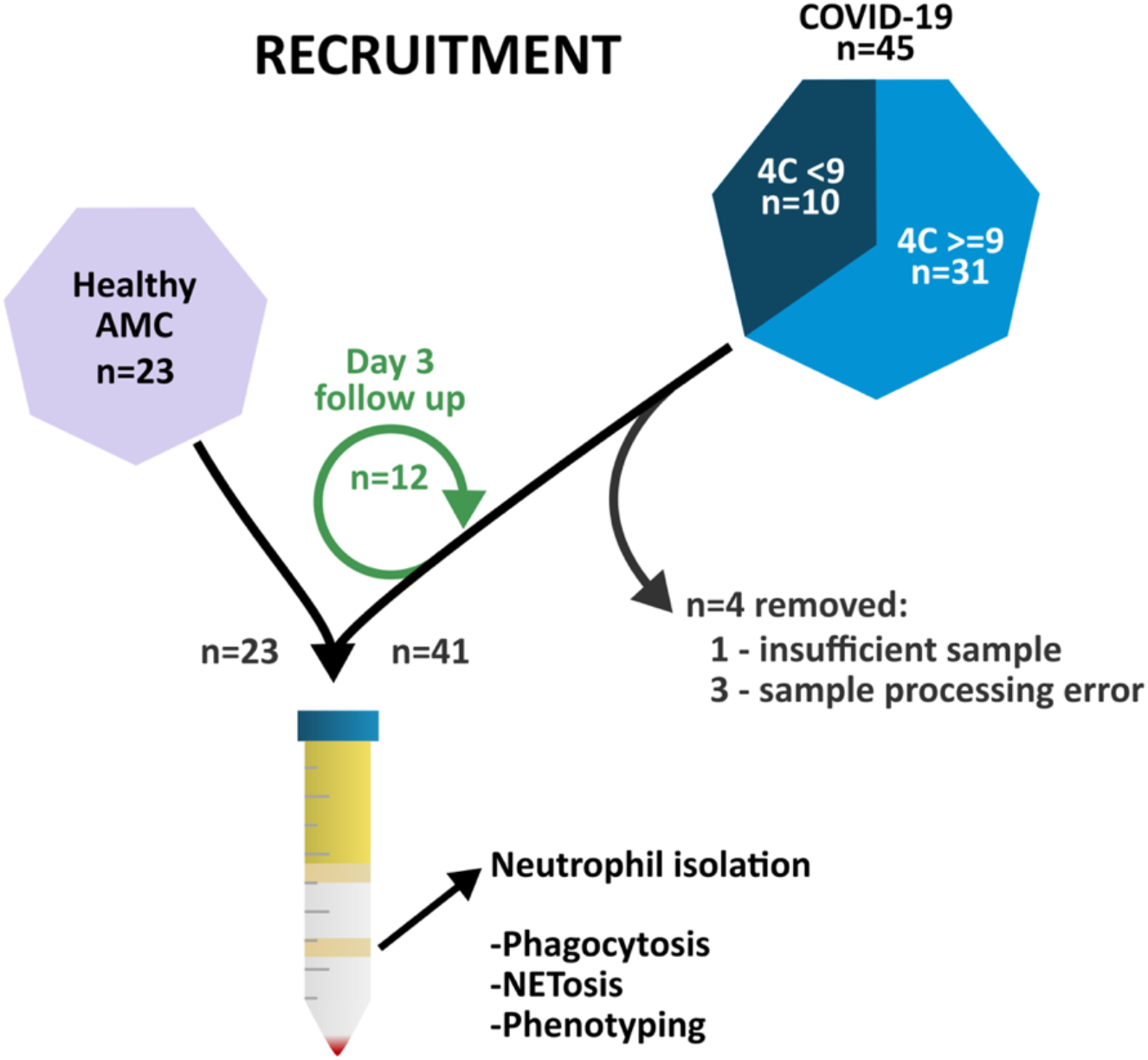
Patient recruitment. 45 patients with COVID-19 were recruited from the Queen Elizabeth Hospital Birmingham, alongside 23 age matched controls. Blood was taken, and neutrophils isolated.

Patients were recruited within 48 hours of hospital admission due to pneumonitis/pneumonia related to COVID-19. All patients had a positive COVID-19 PCR swab. No patients received novel treatments or were part of a COVID clinical medicinal trial on recruitment to this study. Exclusion criteria were admission outside timeframe, no evidence of pneumonia/pneumonitis, long-term oral steroid use, haematological malignancy causing neutropenia, any cause of immunosuppression, recruitment into a trial for novel therapy, admission to ITU, palliative or imminent treatment withdrawal within 24 hours and pregnancy. Patients with COPD or asthma were excluded due to known alterations in neutrophil function in these patient groups [29]. COVID-19 patients were stratified using the 4C Mortality Score for COVID-19, separated by scores <9 and >=9 (in-hospital mortality <9.9% and >31.4% respectively) [30]. Patients were classified with ARDS based on the Berlin criteria; SpO2: FiO2 ratio (SF), converted to PaO2: FiO2 (PF) (SF=57-0.61PF).

Age matched controls (AMC) were either recruited from patients attending pre-booked face-to-face outpatient appointments or from hospital staff. AMC had no evidence of acute illness including COVID-19 within the last two weeks, as assessed by a respiratory physician, and met the other exclusion criteria described above.

Additional details on methodology are provided in the online supplement. These methods include; plasma collection, neutrophil isolation by Pecoll gradient centrifugation, phagocytosis of fluorescent *S. pneumoniae*, NETosis by release of cell-free (cf)DNA, transwell migration, phenotyping by flow cytometry, serum DNase-1 activity assay, plasma cfDNA quantification, citrullinated histone H3 detection and plasma biomarker quantification.

### Statistics

Statistical analysis is described in the online supplement. A power calculation performed on isolated neutrophil NETosis data (80%, alpha 0.05) suggested 18 participants were required in each group (see supplement for details). Data are presented throughout as median (IQR), with each n number representing a separate study participant. Significance was defined at p <0.05. There were no corrections for multiple comparisons but exact p values are given.

## Results

### Clinical characterisation

41 COVID-19 patients (mean age 69.9 years) and 23 healthy AMC (mean age 69.7 years) were included in the study. Demographics are provided in Table 1. COVID-19 patients were admitted to hospital 7 days (range 3-14) after symptom onset and were recruited to the study after median 2 days (range 1-2). Length of hospital stay was 7.7 days (survivors 7.5 days, non-survivors 9.1 days) and the mortality rate was 24% (10/41). 38/41 patients received dexamethasone as part of their acute treatment as per standard of care [31]. 17/41 patients had ARDS as defined by the Berlin criteria with the exception of ventilation pressure [32], and of these 8 had moderate to severe ARDS.

**Table 1.**
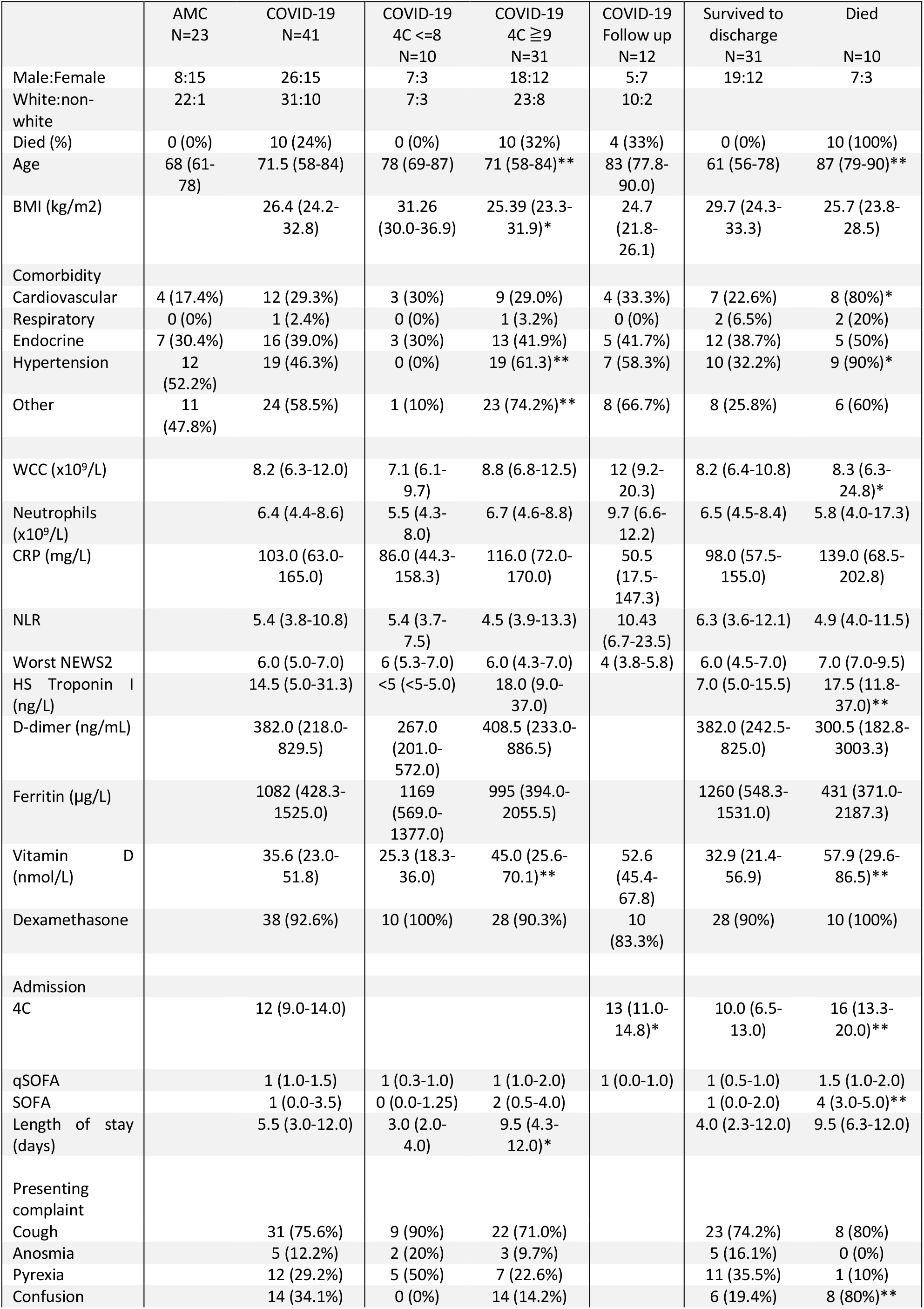
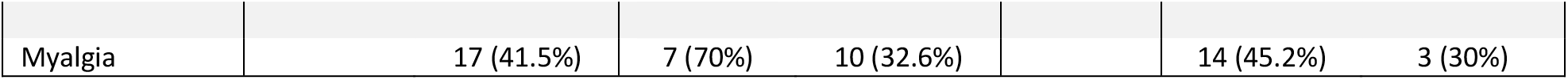
Patient demographic details. Data was collected at the time of recruitment. ± values indicate SD. WCC-White cell count; CRP-C-reactive protein; NLR-neutrophil: lymphocyte ratio; NEWS2-National Early Warning Score2; HS Troponin I-high sensitivity troponin I; 4C-4C Mortality Score for COVID-19; qSOFA-quick Sepsis-related Organ Failure Assessment; SOFA-Sepsis-related Organ Failure Assessment. *<0.05, **< 0.001, *** <0.0001, normally distributed results shown with mean ±SEM, not normally distributed results shown with median (IQR Q1-Q3). Other diagnoses were included if they constituted significant impact on quality of life or regular medication, included but not limited to: severe peripheral vascular disease with active ulceration, dementia, chronic kidney disease, stroke, rheumatoid arthritis, childhood polio, diverticulosis, obesity, alcohol liver disease.

### Neutrophil migration is elevated through a transwell system in COVID-19

Neutrophils from COVID-19 patients demonstrated increased transwell migration towards CXCL-8 compared to AMC (Fold change of neutrophils migrated to CXCL-8 vs. vehicle control: 12.15 (51.4) AMC vs. 40.63 (115.2) COVID-19, p=0.0397, Figure 2A). Migration was not associated with 4C score (p=0.5575, Supplementary Figure 5A), or survival at 28 days (p=0.2563, Supplementary Figure 6A). There was no change in migration at follow up (p=0.1641, Supplementary Figure 4A).

**Figure 2.**
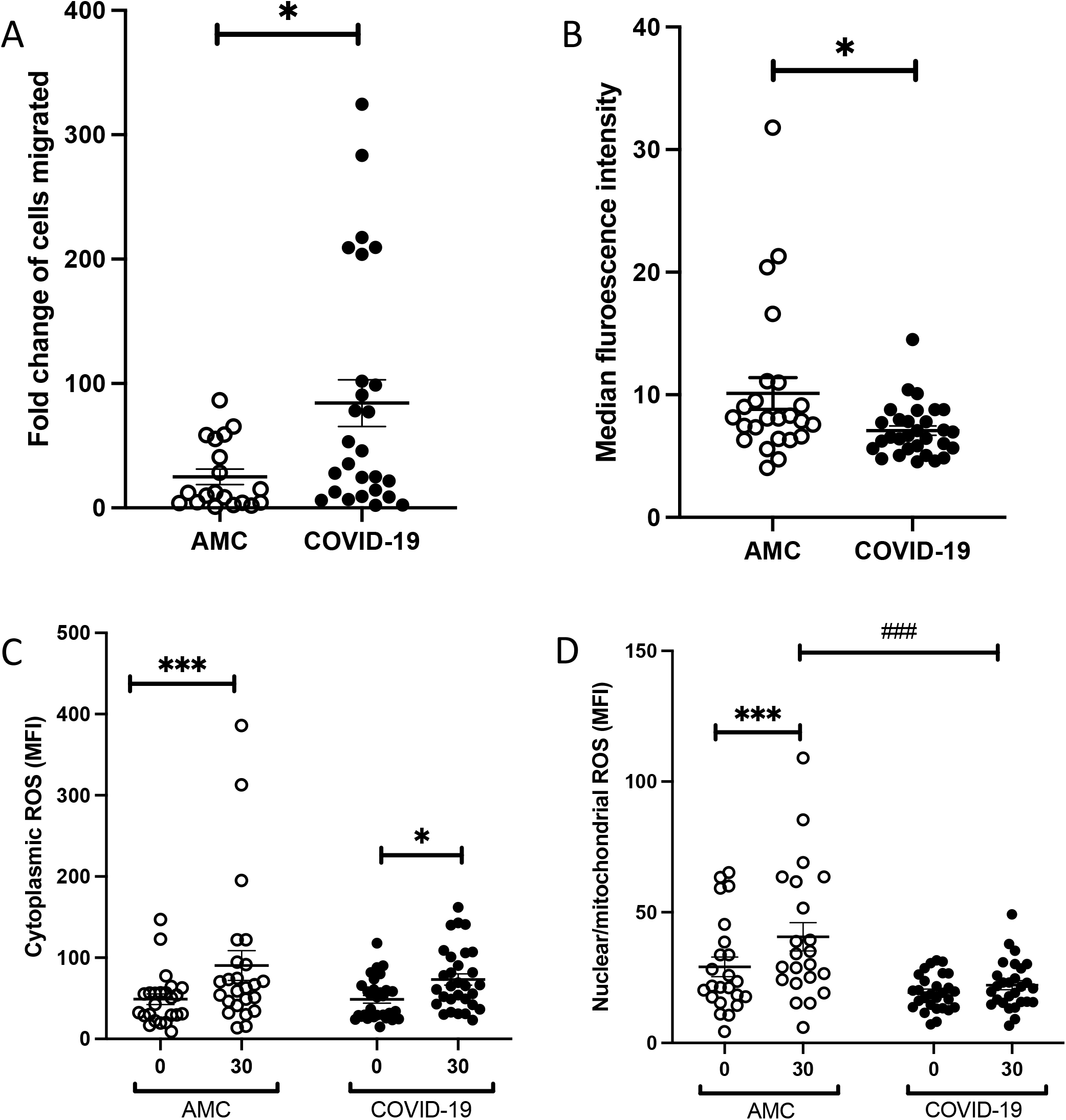
Neutrophil functional responses in COVID-19. **A.** Neutrophil transmigration towards IL-8. Data shows fold change in neutrophils migrated to CXCL-8 compared to vehicle control. AMC, n=18; COVID-19 patients n=28. *p=0.0397, Mann-Whitney U test. **B**. Neutrophil phagocytosis of opsonised S. pneumoniae following a 30-minute incubation. Data shows MFI of positive neutrophils. *p=0.0236 Mann-Whitney U test between AMC (n=24) and COVID-19 patients (n=30). **C**. Cytoplasmic ROS levels measured in neutrophils at baseline (0 minutes) and after phagocytosis. Data were analysed by two-way ANOVA *p<0.0235, ***p=0.0005 between time 0 and 30 minutes. **D**. Nuclear/mitochondrial ROS levels in neutrophils at baseline (0 minutes) and after phagocytosis. Data were analysed by two-way ANOVA and post-hoc test *p<0.0001 between time 0 and 30 minutes. ### p<0.0001 between AMC and COVID-19 patients.

### Neutrophil phagocytosis is impaired in COVID-19

Neutrophil phagocytosis of *S. pneumoniae* was significantly decreased in COVID-19 patients compared to AMC (Median fluorescence intensity (MFI): 8.0 (4.2) AMC vs. 6.6 (2.6) COVID-19, p<0.0236, Figure 2B). Phagocytosis was not associated with 4C score (p=0.3257, Supplementary Figure 5B) or survival at 28 days (p=0.9228, Supplementary Figure 6B). There were no differences in neutrophil viability between patient groups or treatment conditions (p=0.6726, Supplementary Figure 3A).

### Neutrophil derived ROS generation following phagocytosis is impaired in COVID-19

Cytoplasmic (c)ROS, and nuclear/mitochondrial (n/m)ROS were measured in resting neutrophils and following phagocytosis. Compared to unstimulated cells, cROS levels were elevated after phagocytosis in both AMC (MFI: 46.8 (28) baseline vs. 64.9 (51) 30 minutes, p<0.0005) and COVID-19 patients (MFI: 44.1 (35) baseline vs. 65.3 (60) 30 minutes, p=0.0235). However, no differences were observed in cROS levels in either resting (p=0.999) or stimulated (p=0.3946) neutrophils between study groups (Figure 2C).

Compared to resting neutrophils, n/mROS levels were significantly higher after phagocytosis in neutrophils isolated from AMC (MFI: 21.8 (21) baseline vs. 32.0 (38), p<0.0001), but not in COVID-19 patients (MFI: 18.9 (12) baseline vs. 21.2 (12), p=0.0610). No significant differences were found in the levels of n/mROS in resting neutrophils (MFI: 21.8 (21) AMC vs. 18.9 (12) COVID-19, p=0.0610). However, reduced levels of n/mROS were measured in neutrophils isolated from COVID-19 patients following phagocytosis when compared to AMC (MFI: 32.0 (38) AMC vs. 21.2 (12) COVID-19, p<0.0001, Figure 2D).

### Neutrophil NETosis is elevated in COVID-19

Higher levels of cfDNA were detected in supernatants obtained from COVID-19 patient neutrophils post-PMA treatment compared to AMC (fold change in absorbance of neutrophils stimulated with PMA vs. vehicle control: 1.29 (0.32) AMC vs. 1.53 (1.66) COVID-19, p=0.0366, Figure 3A). There were no differences in resting neutrophils NET production between COVID patients and AMC (Supplementary Figure S3). When COVID-19 patients were stratified by disease severity, more severe disease was associated with increased NETosis (fold change: 1.17 (0.35) Low 4C vs. 1.41 (0.80) High 4C, p=0.0118, Figure 3B.

**Figure 3.**
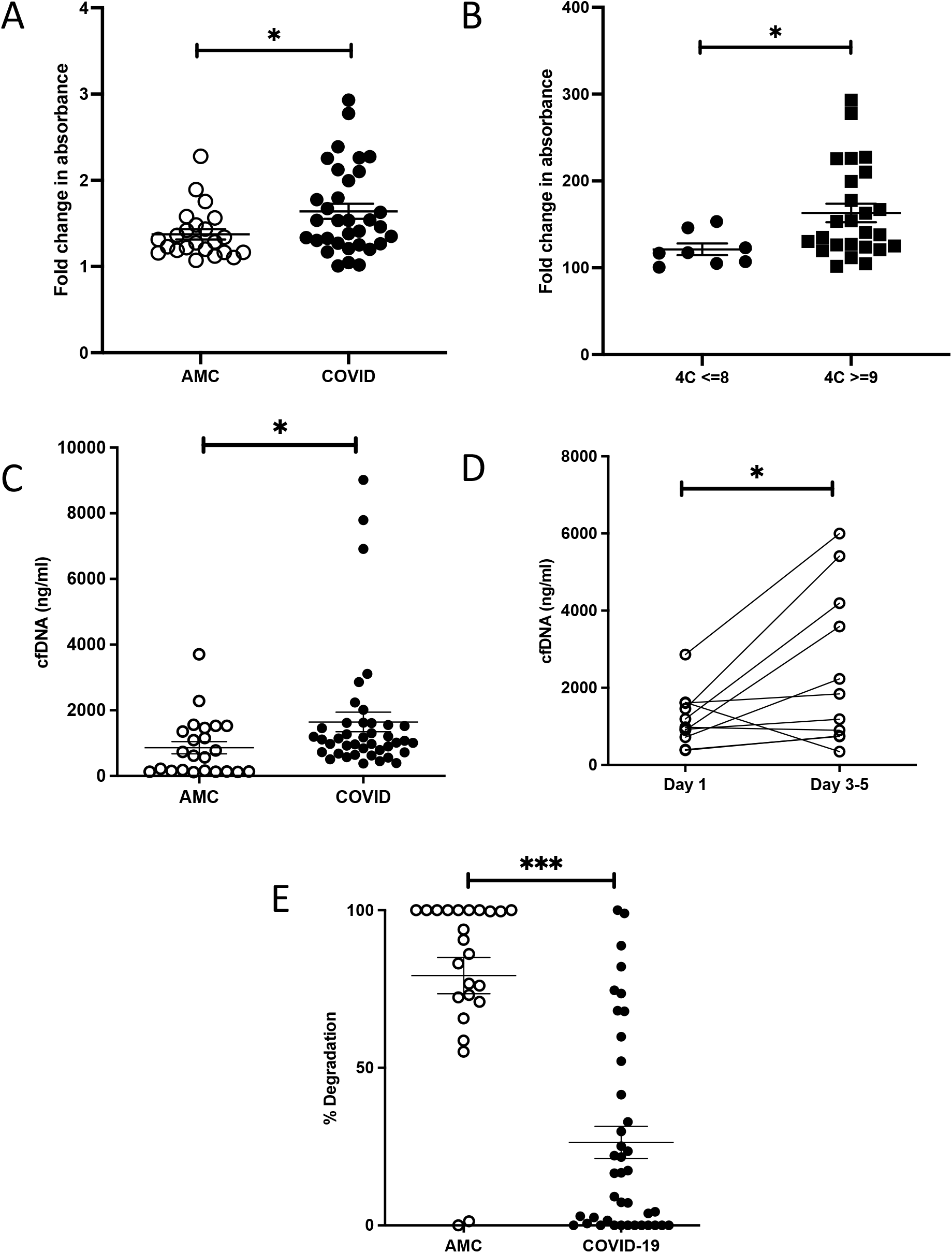
NETosis in COVID-19. **A.** NET release by neutrophils stimulated with PMA for 3 hours, measured as absorbance of cfDNA stained with Sytox green. Data shows fold change in absorbance of PMA stimulated neutrophils compared to vehicle control. Data analysed by Mann-Whitney U test between AMC (n=23) and COVID-19 patients (n=33), *p=0.0366. **B**. Comparison of NET release by activated neutrophils in patients with low 4C (n=8) or high 4C (N=26) score. Data analysed by Mann-Whitney U test, *p=0.0118. **C**. Plasma cell free DNA levels measured by flurometry in AMC (n=23) and COVID-19 patients (n-41). Data analysed by Mann-Whitney U test, *p=0.0153. **D**. Comparison of cfDNA levels measured by flurometry in COVID-19 (N=10) on day 1 and on day 3-5. Data analysed by Wilcoxon test. *p=0.0186. **E**. % DNA degradation by serum NETs in AMC (n=23) and COVID-19 (n=41). Data analysed by Mann-Whitney U test ***p<0.0001.

At hospital admission, COVID-19 patients presented with higher concentrations of plasma cfDNA compared to AMC (621ng/ml (1324) AMC vs. 1071ng/ml (856) COVID-19, p=0.0153, Figure3C), which persisted at Day 3-5 day (p=0.0186, Figure. 3D).

To determine whether neutrophils, via NETosis, were a source of the cfDNA, plasma samples were screened for the presence of CitH3, a protein that decorates the DNA backbone of NETs [33]. Western blotting revealed the presence of CitH3 in 6 out of 8 samples analysed (75%, Supplementary Figure 7).

Using NETs as a substrate, serum DNase activity was lower in COVID-19 patients at the time of hospital admission when compared to AMCs (% degradation: 88.4% (93) AMC vs. 12.8% (49), p<0.0001, Figure 3E).

### Neutrophil phenotype is altered in COVID-19

To determine whether changes observed in neutrophil function were associated with phenotype, surface expression of key surface molecules were investigated by flow cytometry. A table of percentage receptor expression and MFI is shown in table 2.

**Table 2.** Percentage of cells expressing, or MFI of isolated neutrophil cell surface markers measured by flow cytometry in AMC (n=22) or COVID-19 patients (n=34). Data displayed as median (IQR). Data analysed by individual Mann-Whitney test between AMC and COVID-19, with significance shown in bold.

| Receptor | % expression |  |  | MFI |  |  |
| --- | --- | --- | --- | --- | --- | --- |
|  | AMC<br>N=22 | COVID<br>N=34 | P value | AMC<br>N=22 | COVID<br>N=34 | P value |
| CD10 | 95.1<br>(7.72) | 95.3<br>(13.6) | 0.977 | 669 (697) | 836 (518) | 0.623 |
| CD11b | 81.0 (8.9) | 67.8<br>(43.9) | **\*0.0229** | 343 (119) | 40 (223) | **\*0.005** |
| CD54 | 33.4<br>(35.3) | 71.3<br>(21.2) | **\*$<0.0001$** | 21 (38) | 73 (58) | **\*0.008** |
| CD62L | 19.8<br>(14.7) | 31.0<br>(15.3) | **\*0.0096** | 62 (46) | 94 (121) | 0.379 |
| CXCR2 | 100 (0.1) | 100 (0.2) | 0.119 | 2472<br>(1632) | 2031<br>(1600) | 0.153 |
| CXCR4 | 96.6<br>(11.3) | 96.7<br>(9.07) | 0.508 | 2216<br>(4130) | 1724<br>(4177) | 0.552 |
| CD66b | 99.8 (0.5) | 99.8 (0.3) | 0.770 | 257 (213) | 186 (161) | 0.353 |
| CD11c | 99.7<br>(1.65) | 99.9<br>(1.05) | 0.101 | 220<br>(1274) | 1148<br>(670) | **\*0.008** |
| PD-L1 | 99.6<br>(40.4) | 98 (27.9) | 0.452 | 157 (166) | 70 (132) | 0.093 |

Both the percentage of neutrophils expressing the activation marker CD11b (81% (8.9) AMC vs. 67.8% (43.9) COVID-19, p=0.0229), and its surface expression (MFI:343 (119) AMC vs. 40 (141) COVID-19, p=0.0003, Figure 4A) were reduced in COVID-19 patients. Expression of the adhesion molecule CD11c was also elevated in COVID-19 patients (MFI: 220 (1274) AMC vs. 1148 (670) COVID-19, p<0.0008, Figure 4B).

**Figure 4.**
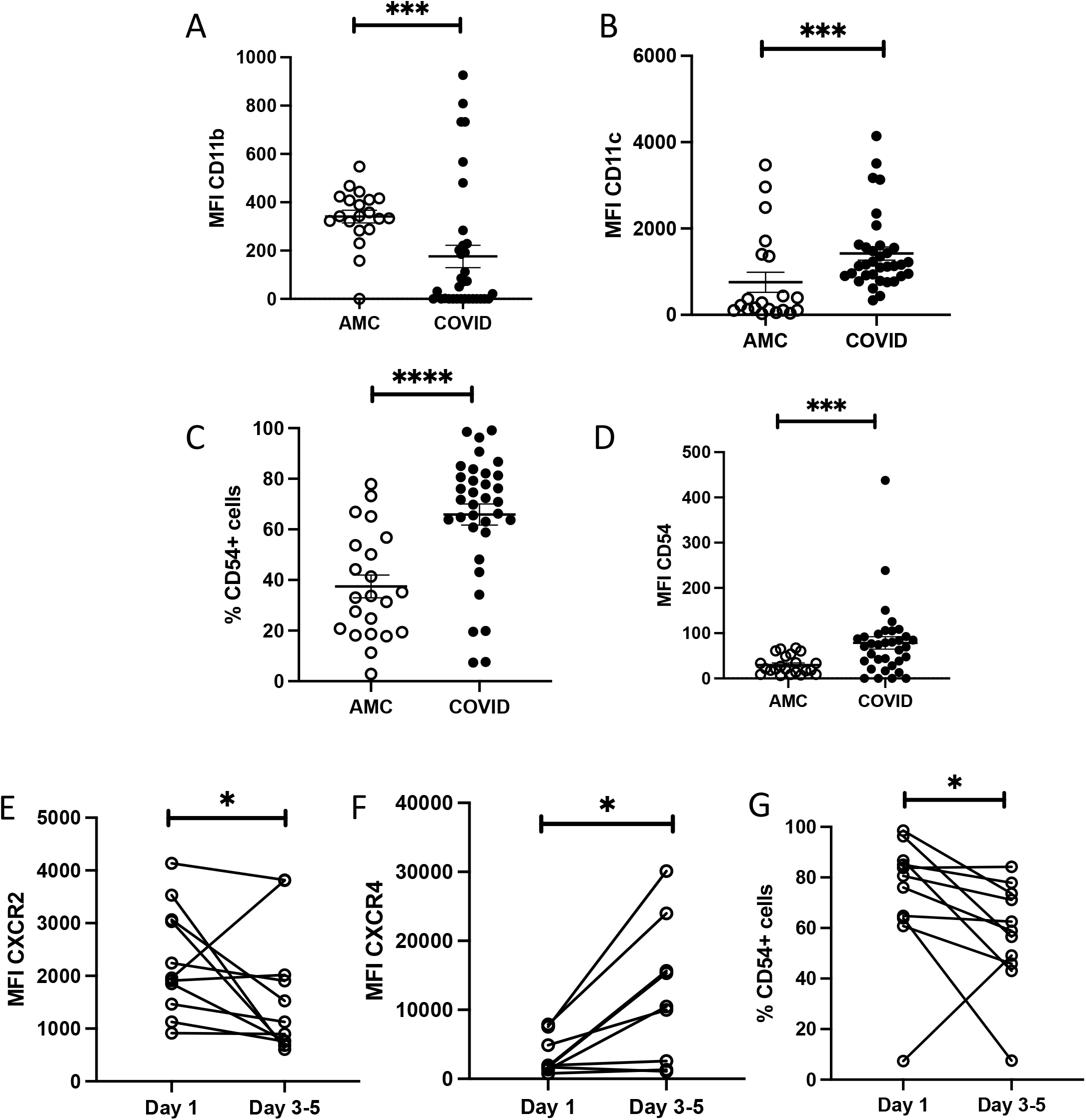
Neutrophil phenotype in COVID-19. Expression of cell surface markers on neutrophils measured by flow cytometry. Data shows MFI of CD11b (**A**, p=0.0003)), CD11c (**B**, p=0.0008), % expression of CD54 (**C**, p=<0.0001) andMFI of CD54 (**D**, p=0.0008) in AMC (n=22) and COVID-19 patients (n=34). Data were analysed by Mann-Whitney U test. Expression of CXCR2 (**E**, p=0.0322), CXCR4 (**F**, p=0.0273) and % expression of CD54 (**G**, p=0.0420) on COVID-19 neutrophils on day 1 or follow up (day 3-5). Data are expressed as individual points, analyzed by Wilcoxon test (n=11).

Both the percentage of neutrophils expressing CD54, a marker of reverse transmigration (33.4% (35.3) AMC vs. 71.3% (21.2) COVID-19, p<0.0001, Figure 4C) and its surface expression (MFI: 21 (38) AMC vs. 73 (58) COVID-19, p=0.0008 Figure 4D) were elevated in COVID-19 patients.

Expression of CD66b, CD62L, CD10, CXCR2, CXCR4 and PD-L1 did not differ between AMC and COVID-19 patients (see table 2). There was no association of neutrophil phenotypic marker expression with 4C severity score, or in survivors and non-survivors.

On day 3-5 follow-up, relative to baseline readings, there was a decrease in CXCR2 expression (MFI: 1960 (1601) Day 1 vs. 1126 (1262) Day 3-5, p=0.0322, Figure 4E) and a significant increase in CXCR4 expression (MFI: 1785 (3951) Day 1 vs. 10248 (16483) Day 3-5, p=0.0273, Figure 4F). The percentage of cells expressing CD54 was significantly reduced at 3-5 day follow up (80.6% (34.6) Day 1 vs. 58.9% (38.3) Day 3-5, p=0.0420, Figure4G)

### Systemic inflammatory mediators are elevated in COVID-19

COVID-19 patients showed elevated levels of IL-6 (p<0.009, Figure 5A), VEGF (p<0.0001, Figure5B), sTNFRI (p<0.0001, Figure 5C), and MPO (p<0.0001, Figure 5D), but not GM-CSF (p=0.3375, Figure 5E) when compared to AMC.

**Figure 5.**
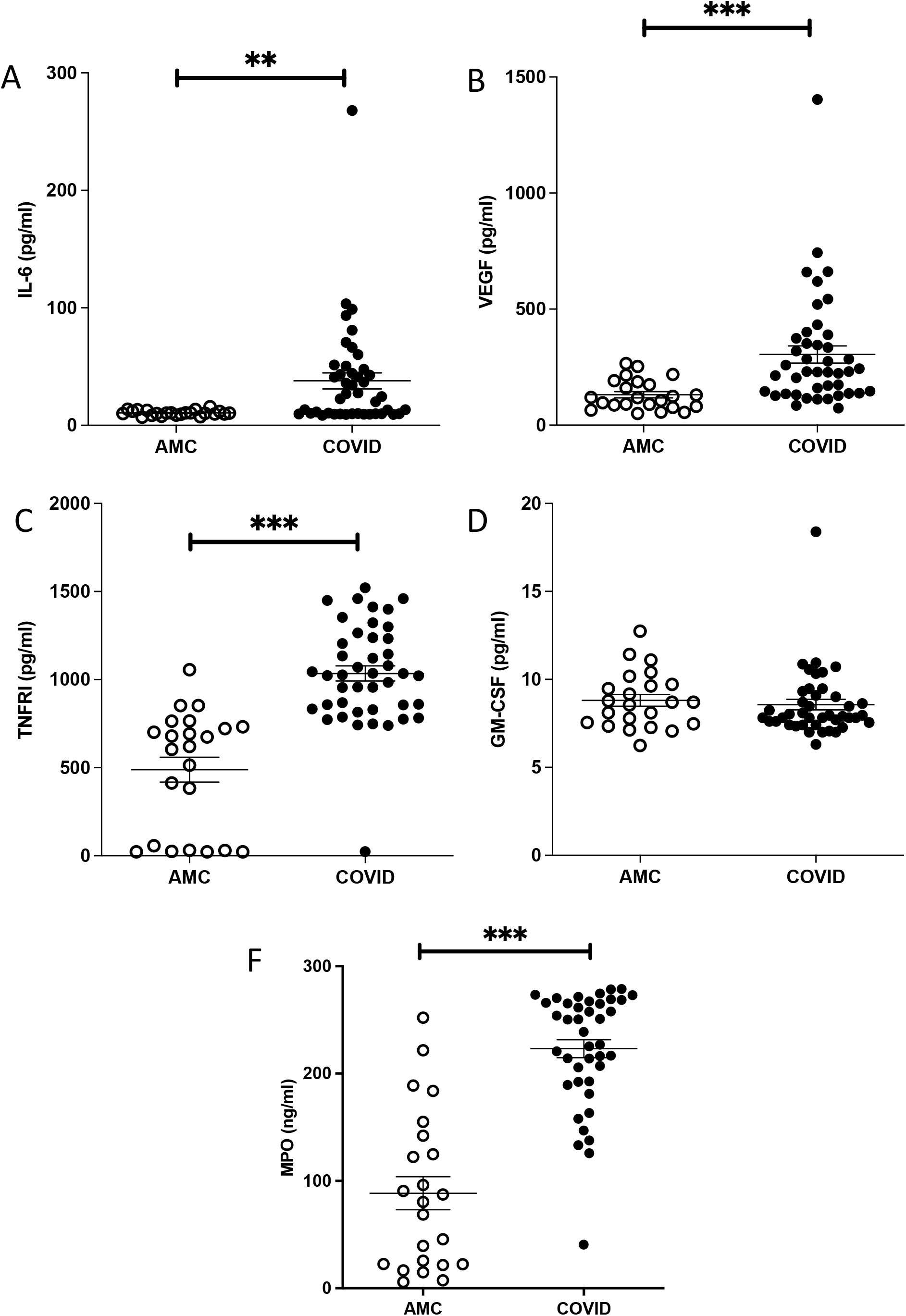
Plasma cytokine levels in COVID-19. Plasma concentrations of **A**. IL-6 (**A**, p=0.009), VEGF (**B**, p<0.0001), sTNFRI (**C**, p<0.0001), GM-CSF (**D**, p=0.3375) and MPO (**E**, p<0.0001) in AMC (n=24) and COVID-19 patients (N=41). Data analysed by Mann-Whitney U test.

Only 2/41 COVID-19 patients demonstrated a hyperinflammatory phenotype when stratified using the algorithm developed to classify non-COVID ARDS phenotypes[34].

IL-6 and sTNFRI concentrations were higher in non-survivors versus survivors (p=0.0027, p=0.0420 respectively, Supplementary Figure. 6) and raised in those patients with a 4C score >= 9 (IL-6 p=0.0059, sTNFRI p=0.0478, Supplementary Figure. 5).

IL-6 levels were significantly raised in patients with moderate to severe ARDS compared to mild (p=0.0468, Supplementary Figure. 6). We observed no associations between the severity of ARDS and VEGF (p=0.1164) or sTNFRI (p=0.3953).

## Discussion

We present novel findings of COVID-associated neutrophil dysfunction, across all main effector functions. In summary, compared to AMC, systemic neutrophils from patients hospitalised with moderate severity COVID-19 demonstrated increased migration, impaired anti-microbial responses including reduced phagocytosis and nuclear/mitochondrial ROS generation after phagocytosis. Later/end phase neutrophil responses were increased, namely *ex-vivo* NETosis with evidence of increased systemic NETosis, coupled with reduced DNase activity. We also show an altered but distinct neutrophil phenotype, not compatible with a purely activated, immature, senescent or anti-inflammatory phenotype as described before (summarised in Figure 6). Our data suggests the energetics of the cells were not overtly compromised, as some “high energy-consuming” functions (such as migration [35]) were elevated. Of note, some of these changes have been described by authors studying COVID-19 before the widespread use of dexamethasone as standard of care [27, 36], suggesting our results are not a treatment effect.

**Figure 6.**
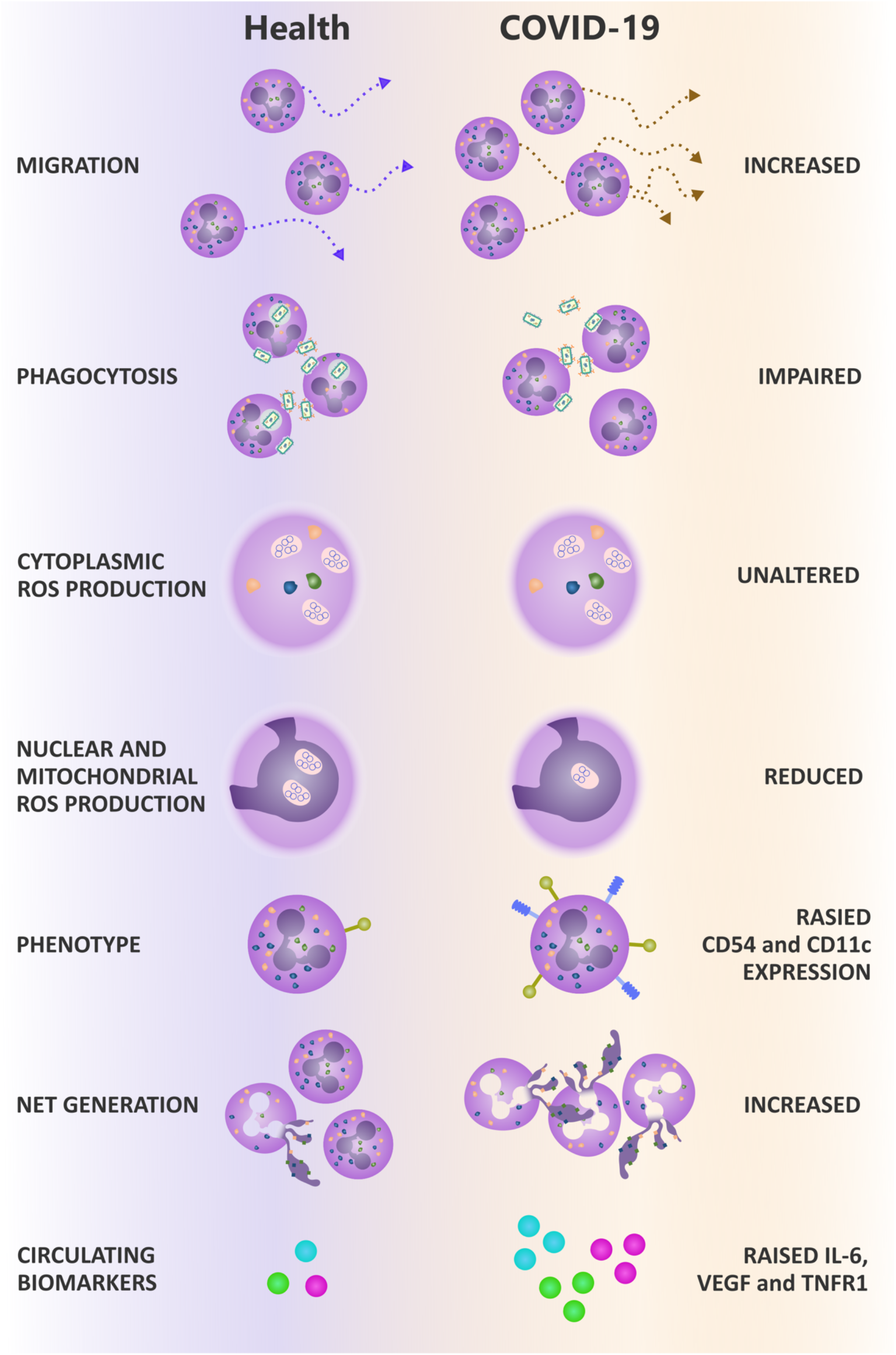
Summary of results. Compared to AMC, neutrophils isolated from non-ICU COVID-19 patients demonstrate increased migration, impaired phagocytosis and reduced nuclear/mitochondrial ROS generation. COVID-19 neutrophils have altered phenotype, displaying increased expression of migration markers CD54 and CD11c. COVID-19 patients also display elevated NETosis, both ex vivo and in the circulation, and elevated pro-inflammatory cytokines. This contributes to disease pathogenesis in COVID-19.

Individually, as described in other studies, these changes to effector function could compromise aspects of host defence. Collectively, these changes represent a clear mechanism for significant tissue damage. Poor phagocytosis would impede pathogen clearance, increasing the likelihood of secondary infection and amplifying inflammation. NETosis is implicated in tissue damage and thrombotic events in several disease settings [37, 38]. The inability to clear NETs through reduced DNAse activity would further augment NETosis-associated tissue damage [39].

Secondary infection in COVID-19 is associated with increased severity of lung disease and poorer outcomes [40, 41]. Impaired neutrophil antimicrobial responses towards *S. pneumoniae*, the most common bacteria implicated in secondary infection in COVID-19 [42], alongside impaired intracellular ROS generation important for phagosomal bacterial killing [43], may contribute to the incidence of secondary infection and poorer outcomes for these patients.

Elevated NETosis [23, 24, 44] and increased systemic concentrations of cfDNA [23, 24] have been described previously in small numbers of COVID-19 patients. Our observation of reduced serum DNase activity builds on studies showing reduced plasma concentrations of Gelosin in COVID-19; Gelosin depolymerises filamentous actin, an inhibitor of DNAse activity [45-47]. Thus, a circulating microenvironment dominated by negative regulators of DNase-1 could offer a potential mechanistic explanation for the impaired DNase-1 activity we report, with elevated NETosis contributing to host tissue damage and thrombotic events.

The collective pattern of neutrophil dysfunction in COVID-19 speaks of alterations to mechanosensing within these cells. Elevated migration and impaired phagocytosis could both be linked to reduced pseudopod extrusion, which is known to increase migratory speed [48]. Also, pseudopods are involved in phagocytosis, with reorganisation of the cell composition to enable bacterial engulfment [49]. Phosphoinositide 3-kinase (PI3K) is a key intracellular signalling molecule involved in chemotaxis, cytoskeletal rearrangement for phagocytosis, and superoxide generation [50] and has most recently been implicated in NETosis [51]. Aberrant PI3K signalling is linked to increasing age, and PI3K inhibitors have been show to improve neutrophil migratory accuracy in the elderly [9], while reducing NET formation *ex vivo* [52, 53]. This suggests there might be tractable targets across neutrophil cellular functions, which could be manipulated to enhance some responses (phagocytosis) while reducing others (NETosis), and to improve innate immune responses in COVID-19.

We observed an altered neutrophil phenotype in moderate COVID-19, not compatible with previously described populations. COVID-19 neutrophils expressed decreased levels of the activation marker CD11b and a lack of CD62L shedding, which has previously been observed in sepsis [54]. We saw elevated expression of the immunosuppressive marker CD11c, also elevated in sepsis [54], but no changes in the expression of CD10; a marker of immature neutrophils [55], or CXCR2 and CXCR4; markers of senescence [56]. This contrasted with RNAseq studies that reported populations of immature and senescent neutrophils in severe COVID-19 patients compared to mild patients or non-AMC [22, 57]. Our contrasting data from moderate COVID-19 patients suggests that either the duration of COVID-19 infection or the extreme severity of infection (from not requiring, to requiring organ support) affects cellular dysfunction and, as in pneumonia and sepsis [17], may support a window for therapeutic intervention.

Finally, we observed that COVID-19 neutrophils expressed elevated CD54, a marker of reverse transmigration, whereby neutrophils migrate from the tissues back into the circulation. These cells are capable of high levels of oxidative burst [58], which may contribute to high levels of NETosis. By day 3-5 post-admission, we report increased levels of senescent CXCR4^+^ CXCR2^-^ neutrophils, confirming a report of reduced CXCR2^+^ neutrophils in ICU COVID-19 patients [59]. The majority of COVID-19 hospitalisations and deaths occur in non-ICU wards [28], making this cohort important for targeted intervention. While there was evidence of systemic inflammation, indicated by elevated levels of circulating IL-6, sTNFRI and VEGF, less than 5% of the patients in the current study met the criteria for the hyper-inflammation phenotype described in ARDS [34] and had levels of circulating mediators lower than described in “usual” sepsis [13]. IL-6 and CXCL-8, as well as platelet derived factors and antigen-antibody complexes are thought to drive NET formation, providing another mechanism to link systemic inflammation to neutrophil dysfunction [60-62].

Our data suggests a distinct cellular response in moderate COVID-19 which contribute to on-going immune mediated harm, but which may be modifiable using a targeted therapy given administered at this crucial point in disease progression. While *ex vivo* experiments suggest PI3K inhibition may improve all aspects of neutrophil dysfunction described in the current study [52, 53], this has not tested in COVID-19 patients. The STOP-COVID trial (Superiority Trial Of Protease inhibition in COVID-19, NCT04343898) is assessing the ability of Brensocatib to reduce neutrophil serine protease activity in COVID-19. Brensocatib improves clinical outcomes in bronchiectasis patients by reducing neutrophil proteases systemically [63], and may reduce the consequences of elevated NETosis in COVID-19, described in the current study.

Our data complements studies which showed increased systemic NETosis in COVID-19 [24, 64], and elevates those which show increased NETosis in isolated neutrophils, by including increased patient numbers and appropriate AMC [23, 24, 44]. A study by Masso-Silva *et al*. recently showed elevated neutrophil phagocytosis in sixteen ICU COVID-19 patients compared to non-AMC. Differing results may be due to the differing experimental techniques, disease severity and patient numbers. Their study used polymorphoprep rather than Percoll® to isolate neutrophils, and the authors combined data from multiple blood samples taken over eleven days of hospitalisation. As we show changes in neutrophil phenotype and function over the 3-5 time course of our study, we suggest that combining time points obscures the complex changes occurring in this short-lived cell population. We also used opsonised *S. pneumoniae* for phagocytosis studies, which may be phagocytosed by different mechanisms to *S. aureus* bioparticles [49], confounding results.

### Limitations

This study was limited due to the safety measures required. All experiments were carried out within a BSL2 hood and methods chosen based on tolerance to inactivation/fixation with 4% PFA. Our patients did not include an ICU group, however, mild-moderate disease forms a larger proportion of overall COVID-19 patients, and we believe it is this point in the patient pathway which holds most potential for successful intervention.

### Conclusion

Our study shows that moderate COVID-19 is associated with alterations in neutrophil phenotype, increased migratory capacity and NETosis, and impaired antimicrobial function which contributes to the severity of COVID-19. Elevated NETosis in the lung is associated with disease severity, and elevated systemic NET production is likely to contribute to inflammation, which may drive ARDS associated damage, and thrombosis. Targeting neutrophils and their downstream effectors may be beneficial in the treatment of COVID-19.

## Data Availability

None

## Acknowledgements

JH is supported by the National Institute for Health Research (NIHR) Surgical Reconstruction and Microbiology Research Centre (SRMRC). The views expressed are those of the author(s) and not necessarily those of the NIHR or the Department of Health and Social Care. Special thanks to UK-Coronavirus immunology consortium.

