## Supplementary figure for "Altered neutrophil phenotype and function in non-ICU hospitalised COVID-19 patients correlated with disease severity"

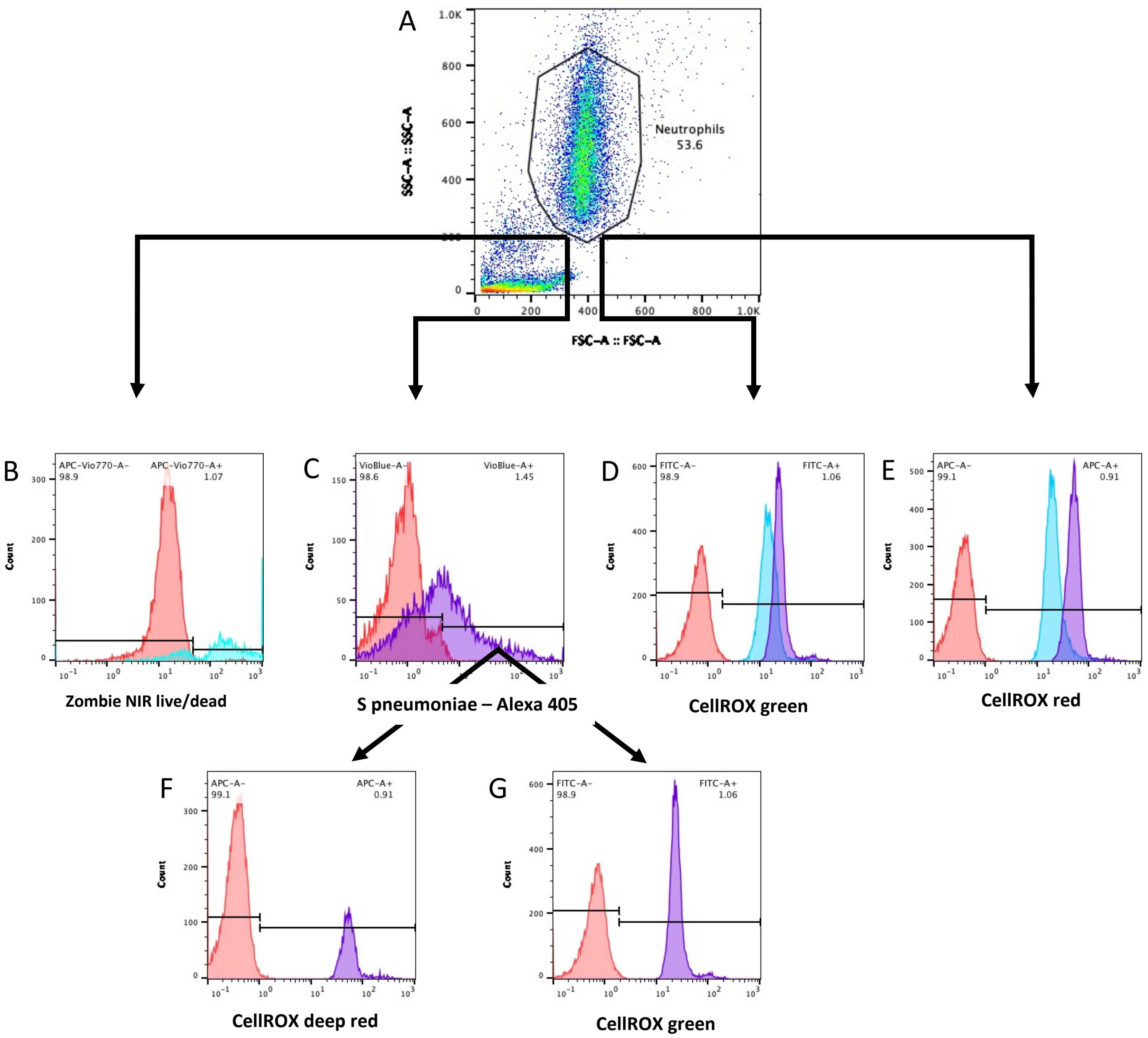


**Supplementary figure 1** – Flow cytometry gating strategy for functional output. Neutrophils were gated on forward vs. side scatter (A). Within this population, multiple parameters were measured: B) Zombie NIR live/dead. C) Phagocytosis of Alexa-405 labelled S. pneumoniae, D) CellROX green, E) CellROX red. Within the SP+ gate, both CellROX deep red (F) and CellROX green (G) were measured.

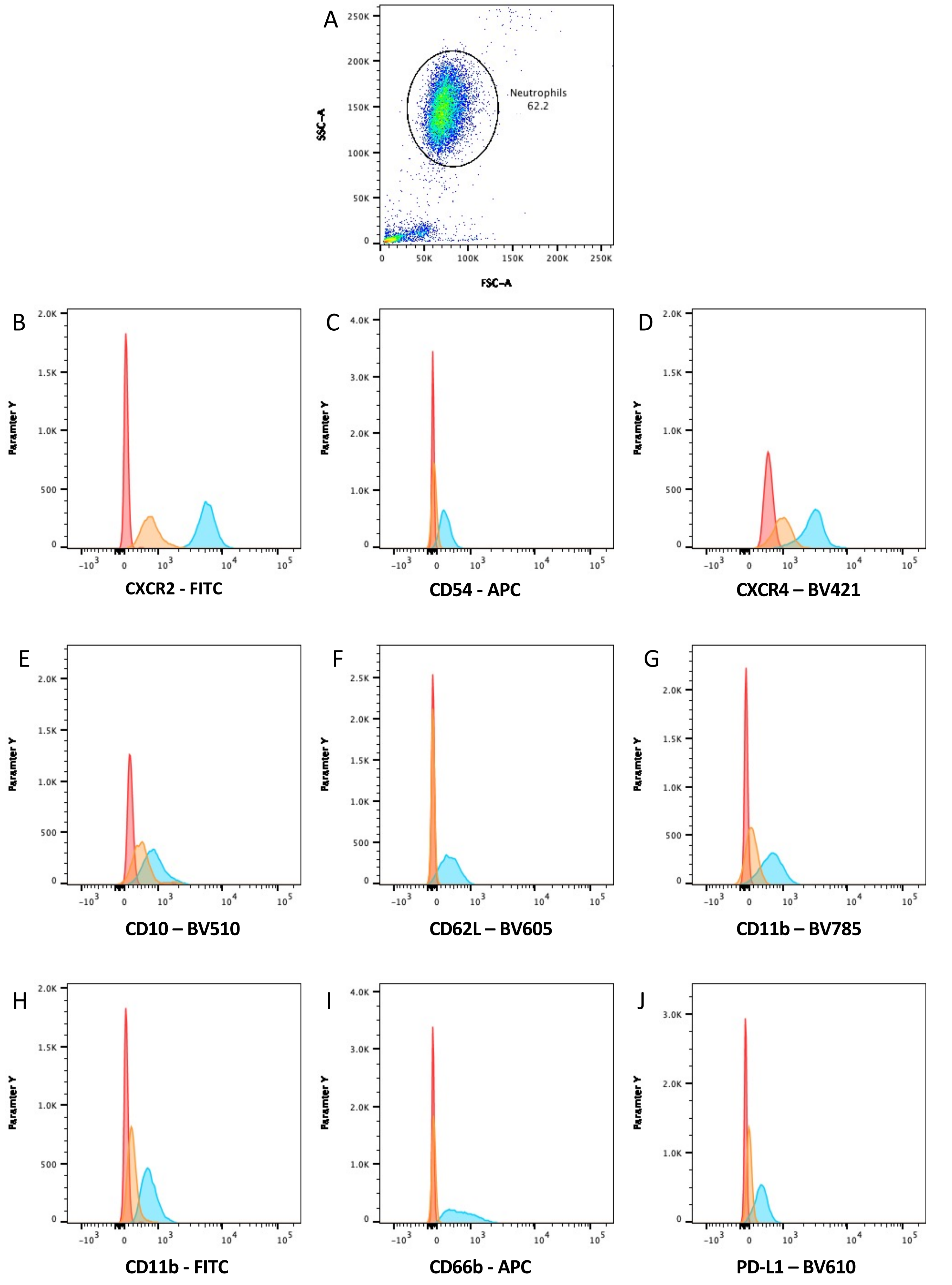


**Supplementary figure 2** – Flow cytometry gating strategy for receptor expression. Neutrophils were gated on forward vs. side scatter. Expression of receptors was analysed within this gate. Histograms show negative unstained control in red, isotype control in orange, and positive sample in blue.

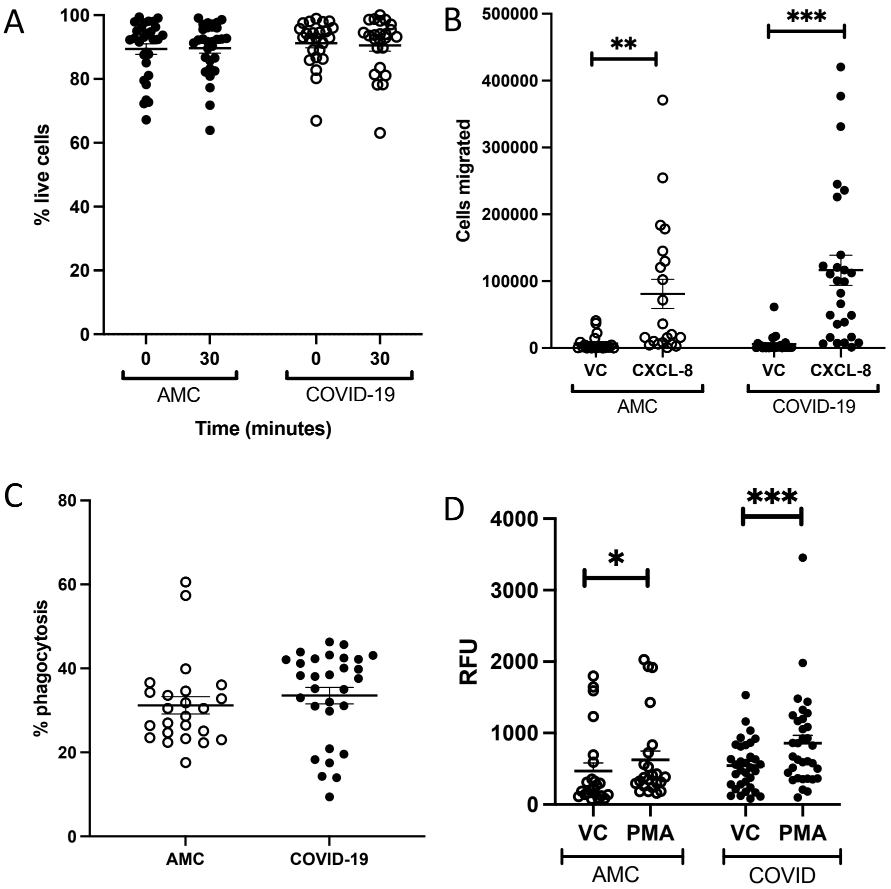


**Supplementary figure 3** – A) Viability of isolated neutrophils at baseline (0) and after phagocytosis (30 minutes) in AMC (n=24) and COVID-19 patients (n=30), measured by Zombie NIR viability stan. Data analysed by two way ANOVA. p=0.6726. B) Number of cells migrated towards CXCL-8 using insall chambers, showing vehicle control (VC) or CXCL-8. AMC n=18, COVID-19 n=28. Analysed by two-way ANOVA. **p<0.0038, ***p<0.0001 VC vs CXCL-8. C) Percentage of neutrophils that phagocytosed *S. pneumoniae* in AMC (n=24) and COVID-19 patients (n=30). p=0.0856*,* analyzed by Mann-Whitney test. D) NETosis by isolated neutrophils, showing absorbance values of cfDNA (relative fluorescence units (RFU), showing vehicle control (VC) or PMA. AMC n=23, COVID-19 n=33. Analysed by two-way ANOVA. **p<0.0321, ***p<0.0001 VC vs PMA.

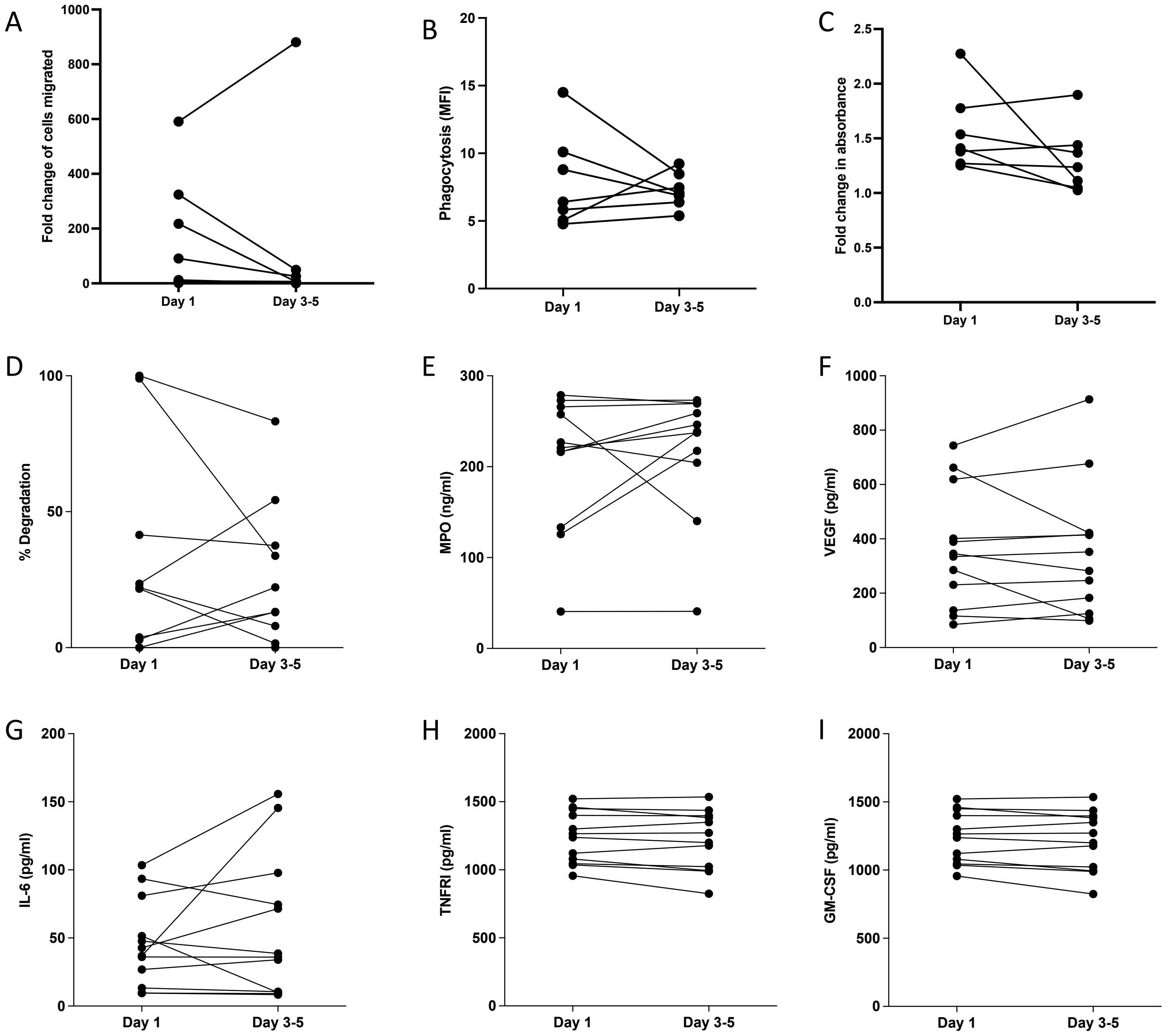


**Supplementary figure 4** – Follow up sample analysis, analysed by Wilcoxon test. A) Neutrophil transmigration towards CXCL-8. Data shows fold change in neutrophils migrated to CXCL-8 compared to vehicle control. N=6, p=0.1641. B) Neutrophil phagocytosis of opsonised S. pneumoniae following a 30-minute incubation. Data shows MFI of positive neutrophils. N=6, p=0.8125. C) NET release by neutrophils stimulated with PMA for 3 hours. Data shows fold change in absorbance of PMA stimulated neutrophils compared to vehicle control. N=7, p=0.1562. D). % DNA degradation by serum NETs, n=10, p=0.7544. E) Plasma MPO levels. N=11, p=0.6063. F) Plasma VEGF levels, n=12, p=0.999. G) Plasma TNFRI levels, n=12, p=0.7125 H) Plasma IL-6 levels, n=12, p=0.9647. I) Plasma GM-CSF levels, n=12, p=0.7125.

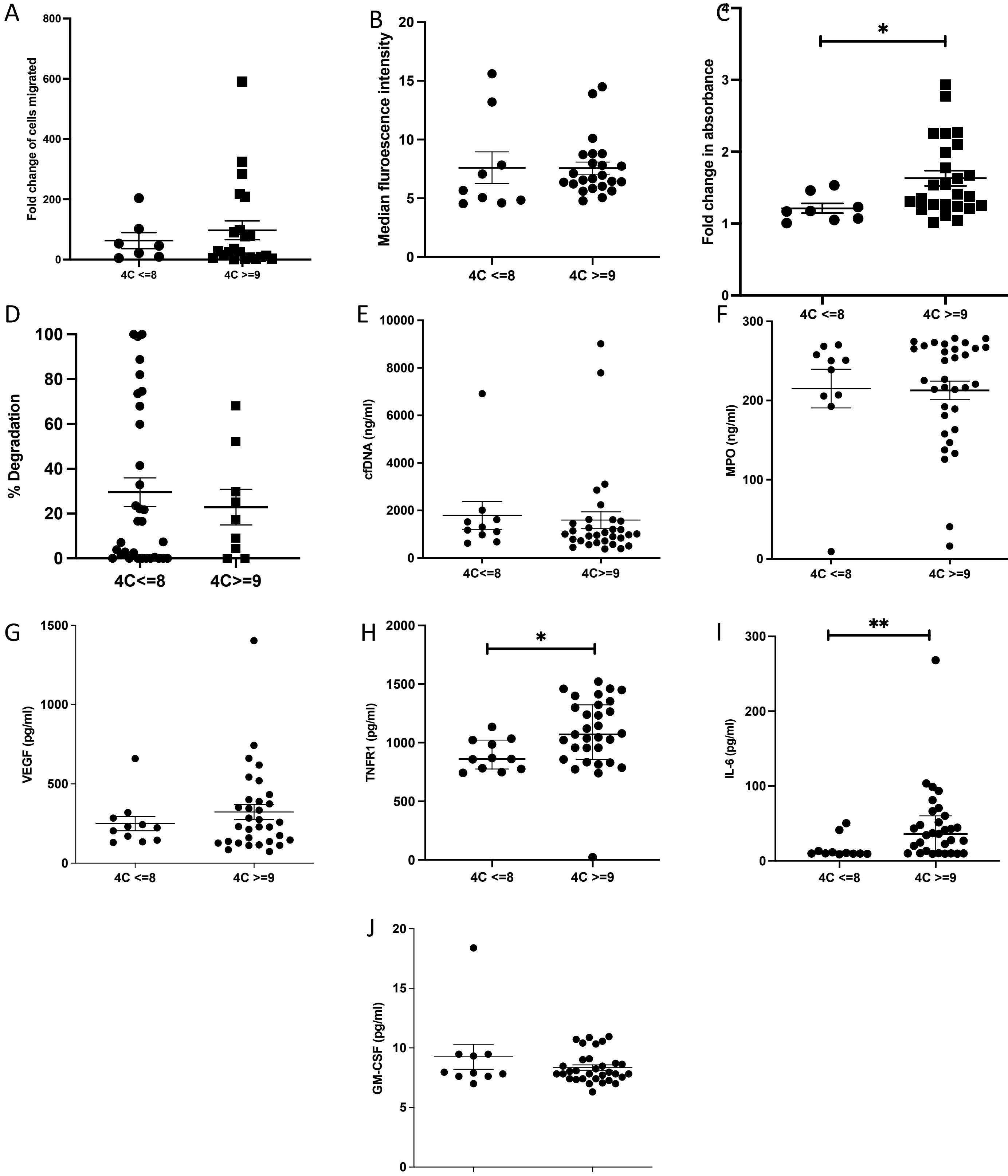


**Supplementary figure 5** – 4C severity score analysis, analysed by Mann-Whitney test. A) Neutrophil transmigration towards CXCL-8. Data shows fold change in neutrophils migrated to CXCL-8 compared to vehicle control. 4C<=8 (n=7), 4C>=9 (n=23), p=0.5575. B) Neutrophil phagocytosis of opsonised S. pneumoniae following a 30-minute incubation. Data shows MFI of positive neutrophils. 4C<=8 (n=9), 4C>=9 (n=23), p=0.3257. C) NET release by neutrophils stimulated with PMA for 3 hours. Data shows fold change in absorbance of PMA stimulated neutrophils compared to vehicle control. 4C<=8 (n=8), 4C>=9 (n=25), *p=0.0118. D). % DNA degradation by serum NETs, 4C<=8 (n=9), 4C>=9 (n=32), p=0.6054. E) Plasma cell free DNA levels, 4C<=8 (n=10), 4C>=9 (n=31), p=0.3001. F) Plasma MPO levels. 4C<=8 (n=9), 4C>=9 (n=32), p=0.9436. G) Plasma VEGF levels, 4C<=8 (n=9), 4C>=9 (n=32), p=0.5961. H) Plasma TNFRI levels, 4C<=8 (n=9), 4C>=9 (n=32), *p=0.0478 I) Plasma IL-6 levels, 4C<=8 (n=9), 4C>=9 (n=32), *p=0.0059. J) Plasma GM-CSF levels, 4C<=8 (n=9), 4C>=9 (n=32), p=0.6049.

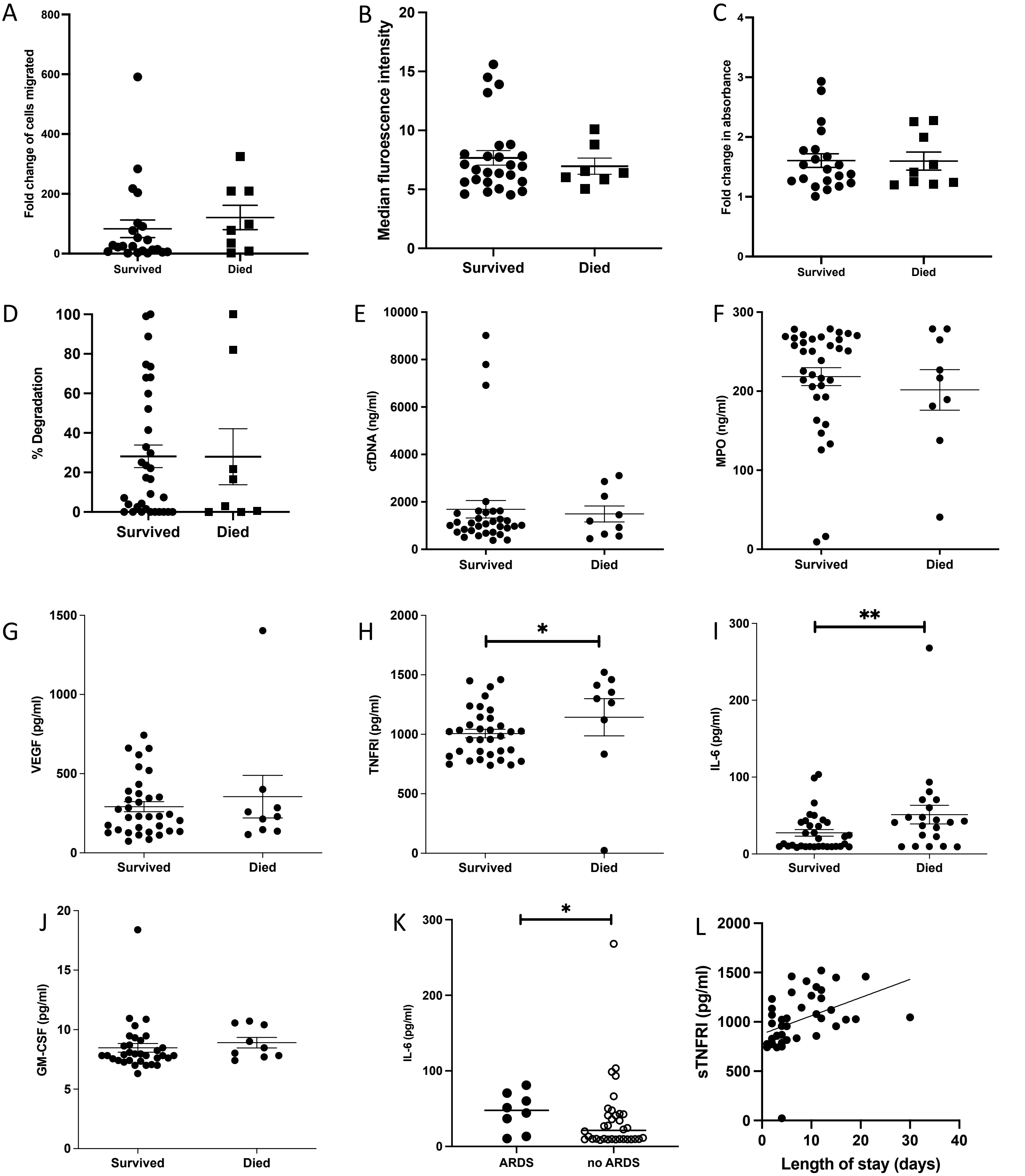


**Supplementary figure 6** – Survival analysis, analysed by Mann-Whitney test. A) Neutrophil transmigration towards IL-8. Data shows fold change in neutrophils migrated to CXCL-8 compared to vehicle control. Survived (n=23), died (n=8), p=0.2563. B) Neutrophil phagocytosis of opsonised S. pneumoniae following a 30-minute incubation. Data shows MFI of positive neutrophils. Survived (n=26), died (n=7), p=0.9228. C) NET release by neutrophils stimulated with PMA for 3 hours. Data shows fold change in absorbance of PMA stimulated neutrophils compared to vehicle control. Survived (n=21), died (n=9), p=0.999. D). % DNA degradation by serum NETs, Survived (n=33), died (n=8), p=0.9895. E) Plasma cell free DNA levels, Survived (n=32), died (n=9), p=0.8406. F) Plasma MPO levels. Survived (n=34), died (n=9), p=0.6888. G) Plasma VEGF levels Survived (n=34), died (n=9), p=0.9924. H) Plasma TNFRI levels, Survived (n=34), died (n=9), *p=0.0311 I) Plasma IL-6 levels, Survived (n=34), died (n=9), p=*0.0027. J) Plasma GM-CSF levels, Survived (n=34), died (n=9), p=0.1875. K) Plasma IL-6 levels in patients with ARDS (n=8) or no ARDS (N=34), p=0.0468. L) Correlation of length of hospital day vs. sTNFRI plasma levels. p=0.006, $r^2$=0.1781.

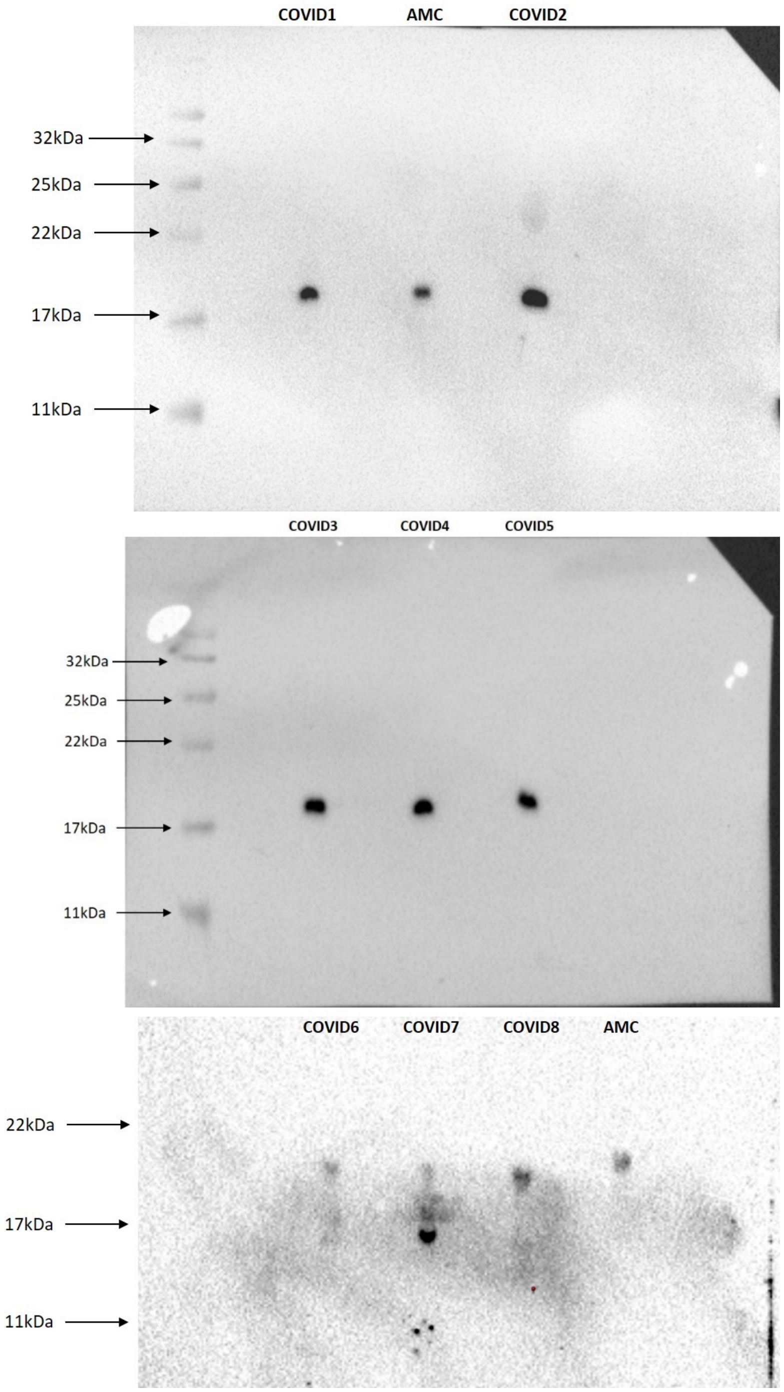


**Supplementary figure 7** – Detection of citrulinated histone 3 (CitH3) in plasma samples from COVID-19 patients and age matched controls. Plasma samples were electrophoresed and blotted for CitH3 (Abcam). Molecular weight markers are indicated with arrows.
