## Supplementary material for "Altered neutrophil phenotype and function in non-ICU hospitalised COVID-19 patients correlated with disease severity": online supplement

**Supplementary text**

**Patients**

Patients were not excluded based on the presence of a 'Do Not Resuscitate' directive. These directives were based on premorbid state and instituted after a multidisciplinary team meeting between parent clinical team, patient and patient family. Some patients were lost to follow up as they were escalated to the intensive care unit for clinical deterioration.

**Preparation of plasma and serum**

For plasma, blood was collected into lithium heparin vacutainers (BD bioscience, UK), then centrifuged at 500g for 10 minutes. For serum, blood was collected into Silica vacutainers (BD Bioscience, UK), allowed to clot for 30 minutes at room temperature (RT), and then centrifuged at 500g for 10 minutes. Plasma or Serum was removed and stored in cryovials at -80°C[1].

**Neutrophil Isolation**

15mls of blood collected in lithium heparin was used for neutrophil isolation by Percol® gradient centrifugation [2]. Neutrophils were counted and diluted to $1x10^6$cells/mL for experimentation as needed. Experiments were prioritised based on cell count, which is reflected in the n for each experiment.

**Phagocytosis**

Serotype 14 *S. pneumoniae* (NCTC 11902, National Collection of Type Cultures) was grown as previously described [3]. Bacteria was heat-killed by incubation at 65°C for 10 minutes as previously described [4]. Bacteria was fluorescently labelled using AlexaFlour 405 NHS ester (Life Technologies) and incubated overnight. Labelled bacteria were washed repeatedly in phosphate buffer saline (PBS) to remove unbound label, resuspended in PBS and stored at -20°C until use. Prior to the experiments, bacteria were opsonised for 1-hour with 10% human serum, then washed by centrifugation. $1.5 \times 10^5$ isolated neutrophils were transferred to FACS tubes, and incubated with bacteria at a ratio of 1600:1 (bacteria:neutrophil), 1μl CellROX green (Thermofischer), 1μl CellROX deep red (ThermoFischer), or 1μl Zombie NIR live/ dead stain (BioLegend) and incubated at 37°C for 30 minutes. Cells were fixed with 4% paraformaldehyde (PFA) for 15 minutes, washed, and resuspended in PBS. Phagocytosis was measured by flow cytometry on a Miltenyi BioTec MACSQuant 10 analyser, with 10,000 neutrophil events recorded for each sample. Flow cytometry results were analysed using FlowJo v10.7.1, with the gating strategy shown in Supp. Figure 1. Phagocytosis results are expressed as either percentage of neutrophil population that were positive for fluorescent bacteria (% phagocytosis), or median fluorescence intensity (MFI) of positive neutrophil population. CellROX green and CellROX deep red data are presented as MFI of neutrophil population.

**NETosis**

$2x10^5$ neutrophils were stimulated with 25 nM phorbol 12-myristate 13-acetate (PMA, Sigma-Aldrich) or vehicle control for 3 hours (37°C, 5% $CO_2$). Post-treatment, 100 µl of supernatant was removed and fixed with an equal volume of 4% PFA for 10 minutes at RT. Supernatants were spun at 2,200 x g for 10 minutes at 4°C to pellet cells, after which 100 µl was transferred to a black 96 well flat-bottomed plate. Samples were stained with 1 µM Sytox green (Life Technologies) for 10 minutes at RT in the dark before fluorescence was measured using a BioTek Synergy 2 fluorometric plate reader (NorthStar Scientific Ltd, UK) with excitation and emission set at 485 nm and 528 nm respectively. Fold change was calculated as:

$$\frac{absorbance\ of\ PMA\ stimulated\ neutrophil\ supernatant}{absorbance\ of\ vehicle\ control\ neutrophil\ supernatent}$$

**Transwell migration**

3 µm transwell inserts (Corning) were placed into 24 well plates containing 600 µl RPMI +/- 100 nM CXCL-8 (Sigma). Neutrophils were resuspended at $1x10^7$ cells/ml in RPMI and $1x10^6$ cells added to each insert. Plates were incubated at 37°C, 5% $CO_2$ for 90 minutes, after which inserts were removed and RPMI supernatants transferred to FACS tubes containing 500 µl 4% PFA. Cells were left for 10 minutes to fix, then centrifuged and resuspended in 600 µl PBS. Cells were counted on Miltenyi Biotech MACSQuant analyser 10, gated on forward vs. side scatter. Fold change was calculated as:

$$\frac{no.\, cells\ migrated\ to\ CXCL-8}{no.\, cells\ migrated\ to\ vehicle\ control}$$

**Neutrophil phenotype by flow cytometry**

Neutrophils ($1x10^6$ cells/ml) were fixed with 4% PFA for 10 minutes, washed, and resuspended in FACS buffer (PBS plus 1% bovine serum albumin (BSA)). 100 µl of cell suspensions were subsequently stained with mouse anti-human monoclonal antibodies or concentration-matched isotype controls for 20 minutes on ice in the dark (Table S1). Flow cytometry was performed on a Fortessa X20 using FACSDiva software, with 10,000 neutrophils recorded for each sample. Our gating strategy is presented in supplementary figure 2. Data are presented as percentage antigen positive cells or median fluorescence intensity, calculated as ($MFI^{sample}$ – $MFI^{isotype\ control}$).

**DNase activity**

NETs served as the substrate for our studies of serum DNase activity. To generate NETs, neutrophils ($5 \times 10^4$) isolated from ethylenediaminetetraacetic acid anti-coagulated blood were dispensed into wells of a 96-well flat-bottomed plate (BD Biosciences) and stimulated for 3 hours (37°C/5% $CO_2$) with 25 nM PMA (Sigma). Post stimulation, NETs were incubated for 6 hours (37°C/5% $CO_2$) with 5% sera diluted in Hank's balanced salt solution supplemented with calcium and magnesium ($HBSS^{+/+}$, Gibco, Life Technologies), or in $HBSS^{+/+}$, alone. Following a 30-minute fixation with 4% PFA (37°C/5% $CO_2$), samples were washed 3 times with PBS prior to a 10-minute incubation at RT in the dark with 1 µM SYTOX green dye (Thermofischer). Fluorescence was measured using a BioTek Synergy 2 fluorometric plate reader (exicitation, 485 nm; emission, 528 nm). NET degradation by 5% serum pooled from 4 healthy control subjects was used to define 100% DNase activity.

**Fluorometric analysis of plasma cell-free deoxyribonucleic acid (CfDNA)**

CfDNA levels in heparinised plasma were measured using a fluorometric based assay. 10 µl of plasma was incubated with 1 µM SYTOX Green dye for 10 minutes in the dark at RT, after which fluorescence was measured using a BioTek Synergy 2 fluorometric plate reader (excitation, 485 nm; emission 528 nm). To determine cfDNA concentrations, a λ-DNA standard curve (Fisher Scientific, UK) was used. Samples were analysed in duplicate with the mean value used to calculate concentrations via extrapolation from the standard curve.

**Detection of citrullinated histone H3 (CitH3) in plasma**

Plasma samples (2 µl), re-suspended in 1X sodium dodecyl sulfate (SDS) buffer, were run on 15% SDS-polyacrylamide gels. Following protein transfer to polyvinylidene difluoride membranes (Bio-Rad, Hertfordshire, UK), blots were probed overnight at 4°C with 1 µg/ml of a rabbit polyclonal antibody against citrullinated histone H3 (ab5103; Abcam, Cambridge, UK). Post incubation, membranes were washed in tris-buffered saline containing 0.001% tween (TBST) and incubated for 1-hour at RT with a goat anti-rabbit secondary antibody conjugated to horse radish peroxidase (HRP; diluted 1:4000 in TBST; GE Healthcare, Buckinghamshire, UK). HRP activity was detected using enhanced chemiluminescence (Bio-Rad). For protein loading, total protein was visualised using a Ponceau S Stain (Sigma-Aldrich).

**Plasma biomarker release**

Biomarkers were measured using BioTechne Quantikine kits for Myeloperoxidase (MPO - DMYE00B), Interleukin (IL)-6 (D6050) and vascular endothelial growth factor (VEGF – DVE00), or Duoset granulocyte macrophage – colony stimulating factor (GM-CSF – DY215-05) according to manufacturer’s instructions.

**Statistics**

Statistical analysis was performed using Prism v9.0.0 (GraphPad Software Inc., San Diego). A Kolmogorov-Smirnov Test was used to determine data distribution. Normally distributed data were analysed using a student’s t-test. A Mann-Whitney U test for unpaired data, or Wilcoxon test for paired data was used to analyse non-normally distributed data. Data are presented throughout as median (IQR), with each n number representing a separate study participant. Significance was defined at $p < 0.05$.

A power calculation performed on isolated neutrophil NETosis data (Group 1 mean=1.374, SD= 0.2876. Group 2 mean= 1.64) with 80% power, and alpha 0.05, revealed 18 participants were required in each group. This was repeated for serum NETosis data (Group 1 mean=26.32 SD= 32.1. Group 2 mean= 79.3) with 80% power, and alpha 0.05, revealed 6 participants were required in each group.

| Receptor | Cat no | Channel | Dilution |
|---|---|---|---|
| CXCR4 | 306518 | BV421 | 1:20 |
| CD10 | 312220 | BV510 | 1:20 |
| CD62L | 304834 | BV605 | 1:100 |
| CD11b | 301346 | BV786 | 1:40 |
| CXCR2 | 320704 | FITC | 1:40 |
| CD54 | 322712 | APC | 1:100 |
| PD-L1 | 329724 | BV605 | 1:100 |
| CD11c | 371516 | FITC | 1:40 |
| CD66b | 305118 | APC | 1:100 |

**Table S1** – Antibodies used for flow cytometry. All antibodies were from BioLegend (UK).

1. Parekh, D., et al., *Vitamin D to Prevent Lung Injury Following Esophagectomy-A Randomized, Placebo-Controlled Trial.* Crit Care Med, 2018. **46**(12): p. e1128-e1135.
2. Sapey, E., et al., *Phosphoinositide 3-kinase inhibition restores neutrophil accuracy in the elderly: toward targeted treatments for immunosenescence.* Blood, 2014. **123**(2): p. 239-248.
3. Dockrell, D.H., et al., *Immune-mediated phagocytosis and killing of Streptococcus pneumoniae are associated with direct and bystander macrophage apoptosis.* The Journal of infectious diseases, 2001. **184**(6): p. 713-22.

4. Bewley, M.A., et al., *Differential Effects of p38, MAPK, PI3K or Rho Kinase Inhibitors on Bacterial Phagocytosis and Efferocytosis by Macrophages in COPD.* PLoS One, 2016. **11**(9).
